# Deep learning-based embryo assessment of static images can reduce the time to live birth in *in vitro* fertilization

**DOI:** 10.1101/2024.10.28.24316259

**Authors:** Lu Yu, Kevin K W Lam, Ernest H Y Ng, William S B Yeung, Lequan Yu, Yin Lau Lee, Yuanhua Huang

## Abstract

The low success rate in *in vitro* fertilization (IVF) may be related to our inability to select embryos with good implantation potential by traditional morphology grading and remains a great challenge to clinical practice. Multiple deep learning-based methods have been introduced to improve embryo selection. However, existing methods only achieve limited prediction power and generally ignore the repeated embryo transfers from one stimulated IVF cycle. To improve the deep learning-based models, we introduce Embryo2live, which assesses the multifaceted qualities of embryos from static images taken under standard inverted microscope, primarily in vision transformer frameworks to integrate global features. We first demonstrated its superior performance in predicting Gardner’s blastocyst grades with up to 9% improvement from the best existing method. We further validated its high capability of supporting transfer learning using the large clinical dataset of the Centre. Remarkably, when applying Embryo2live to the clinical dataset for embryo prioritization, we found it improved the live birth rates of the Top 1 embryo in patients with multiple embryos available for transfer from 23.0% with conventional morphology grading to 71.3% using Embryo2live, reducing the average number of embryo transfers from 2.1 to 1.4 to attain a live birth.

## 1 Introduction

In vitro fertilization (IVF) offers hope to countless couples struggling with infertility. IVF treatment involves ovarian stimulation, oocyte retrieval, and fresh embryo transfer 2-5 days following fertilization in the laboratory. Surplus good quality embryos are cryopreserved by vitrification for transfer later and the survival rate of vitrified embryos after warming can be over 95% [1]. The freeze-all strategy is becoming a preferred routine of IVF with various advantages, including relieving the negative influence of ovarian stimulation [2, 3, 4, 5], improved clinical pregnancy rates and maternal safety [6] and recommended for women with polycystic ovary syndrome or those at risk for ovarian hyperstimulation syndrome [7, 8].

For both fresh and frozen transfers (especially the latter), a key procedure in IVF is the selection of the most viable embryos with the highest implantation potential for transfer, preferably at the blastocyst stage 5 to 6 days following fertilization [9]. Traditionally, embryo selection is primarily based on morphological grading under a microscope by trained embryologists. The most commonly used grading method is the Gardner’s classification, which assesses the degree of expansion, the quality and quantity of the inner cell mass, and trophectoderm of blastocysts [10]. However, the manual morphological grading has to be performed by embryologists with training and years of experience. It is also prone to significant inter- and intra-observer variations, which affect the ability to select the best embryo for transfer and will increase the time to clinical pregnancy, leading to additional mental, physical, and financial stress on patients.

To overcome the limitations of manual morphological embryo grading, efforts have been made to leverage the advances in medical image analysis and develop artificial intelligence (AI) systems to perform automated assessments [11, 12]. Many studies primarily use static embryo images for conventional morphological assessment thanks to the availability and predictive power of such images. The initial applications include automatic grading systems to assist in embryo selection, for example, to predict the Gardner’s grades [13], to apply binary classification between non-blastocysts (classes 1-2) and blastocysts (classes 3-5) [14, 15] or to classify the embryos in three categories via the Veeck and Zaninovic’s system [16]. These prediction targets can generally benefit from access to large training sets of images as they can utilize non-transferred embryos, while the subjective annotations may result in a large variability when directly extending to different IVF clinics and laboratories. Moreover, as the grading system is generally not perfect, predictions based on the Gardner’s classification may have a systematic upper boundary for embryo selection. In contrast, AI may be able to detect more subtle but relevant information from the images directly that can be easily overlooked by human eyes. In most of the studies, a clinical pregnancy is deemed to be the most appropriate measure of embryo viability as it eliminates any confounding patient-related or post-implantation factors, hence providing an effective prediction target [17, 18]. From the patient’s perspective, the live birth outcome is the ultimate goal of any IVF treatment, hence often serving as a critical prediction endpoint, e.g., in [19, 20]. Regardless of the prediction endpoints, it is evident that AI showed excellent power in assisting embryo selection via embryo morphology and clinical outcome prediction, as also summarized in a comprehensive review for AI-assisted IVF research from 2005 to 2022 [12].

Despite many attempts, multiple limitations exist in the current AI systems for embryo selection, both technically and conceptually. First, although efforts have been made to claim AI outperforms embryologists on some evaluation metrics [21] or via simply integrating fresh and post-warmed embryos (regardless of the inherent biases) [20], less attention has been made to establish a more reliable paradigm of equipping embryologists with AI. Second, most current AI systems focus on predicting a single type of clinical results, while multivariate prediction, for example, combining Gardner’s classification, clinical pregnancy, and live birth outcomes, would provide more information for embryo quality assessment and potential treatment. Third, most AI systems only use either fresh or post-warmed static images to perform the prediction, while the two-stage information can be beneficial. Previously, Ahlstrom et al [22] demonstrated that pre-freeze blastocoel expansion, trophectoderm grade, and post-thawed degree of re-expansion are the most significant predictors of live birth in frozen embryo transfer cycles. Such related two-stage (fresh and vitrified-warmed) information on imaging data has not been explored for prediction, partly due to their unavailability in the current clinical routine.

More urgently, an often-overlooked demand is to develop a center-based AI system to perform prioritization among a cohort of multiple embryos from one patient following one stimulated IVF cycle, especially for frozen embryo transfer. From the perspective of application, the intended use of AI for embryo quality assessment is to select the most viable embryo with the highest implantation potential from multiple embryos of a patient for transfer in a stimulated IVF cycle [23]. A recent study [19] showed that the overall performance in predicting live birth could be improved by further adding patient information, such as patient age, previous number of attempts, stimulation protocol, endometrial thickness, and maternal hormone profiles. This is not surprising as the model can adapt to the general trend in success rates with patient information such as maternal age [21], but the increased discrimination performance across all patient groups may not improve ranking ability within individual patient cohorts as the model may be biased towards the patient factors while these factors are usually constant for a certain patient. Therefore, given that the patients’ characteristics cannot be improved, greater emphasis should be placed on selecting the most viable embryos for transfer into the uterus, as this significantly impacts the chances of a successful clinical pregnancy and subsequent live birth.

This paper seeks to contribute to the ongoing dialogue surrounding the potential of AI in optimizing IVF outcomes using static images. We developed a deep-learning-based system Embryo2live with advanced models (mostly in vision transformer architecture) and designed a comprehensive embryo prioritization and quality assessment system for fresh and frozen embryo transfer cycles. The system first aimed to benchmark on a golden-standard grading system for embryologists’ reference to help overcome subjectivity in practice, where our model substantially outperformed the state-of-the-art models as implemented in [9] with higher accuracy. Then, a series of models were deployed to evaluate clinical pregnancy and live birth potentials of both fresh embryos and vitrified-warmed embryos. Uniquely, by integrating both freshand cryopreservedstage information, Embryo2live could reduce the number of embryo transfers and thus shorten the time to clinical pregnancy and live birth for patients. In conclusion, this work empowered embryologists with an effective tool to improve decision-making, reduce subjectivity, increase the chances of clinical pregnancies and live birth per transfer for infertile women undergoing IVF.

## 2 Results

### 2.1 High-level description of the whole framework

In this work, we developed a deep-learning-based system Embryo2live by designing a series of models to assist embryo selection in both fresh and frozen embryo transfer cycles using static images, catering to the workflow of real clinical situations (shown in Fig. 1). In the workflow, embryologists first use Gardner’s grading to assess the quality of blastocysts after 5 or 6 days of culture, capture their static images, and input them into the Embryo2live_fre system to assess embryo quality. Briefly, the Embryo2live_fre system used tailored model architectures for each task to maximize its performance, specifically by using ResNeXt50 [24] for expansion and inner cell mass, ViT-b16 [25] for trophectoderm, clinical pregnancy, and live birth (except ViT-l16 for live birth prediction of frozen embryo transfer samples thanks to the larger sample size; see more details in Methods). This multivariate system can be used for multifaceted purposes. First, the predicted Gardner’s grades can be used as a reference for embryologists’ assessment or to re-examine the potential risk of erroneous prediction. Second, the predicted live birth probability can be used as the primary metric to select the embryo with the highest quality for transfer, despite it may not be ranked first with the conventional grading (as shown in the illustrative example in Fig. 1; top panel). Third, one may consider using the predicted probabilities of both clinical pregnancy and live birth wisely, particularly when the embryo qualities are all suboptimal for a certain patient.

**Figure 1:**
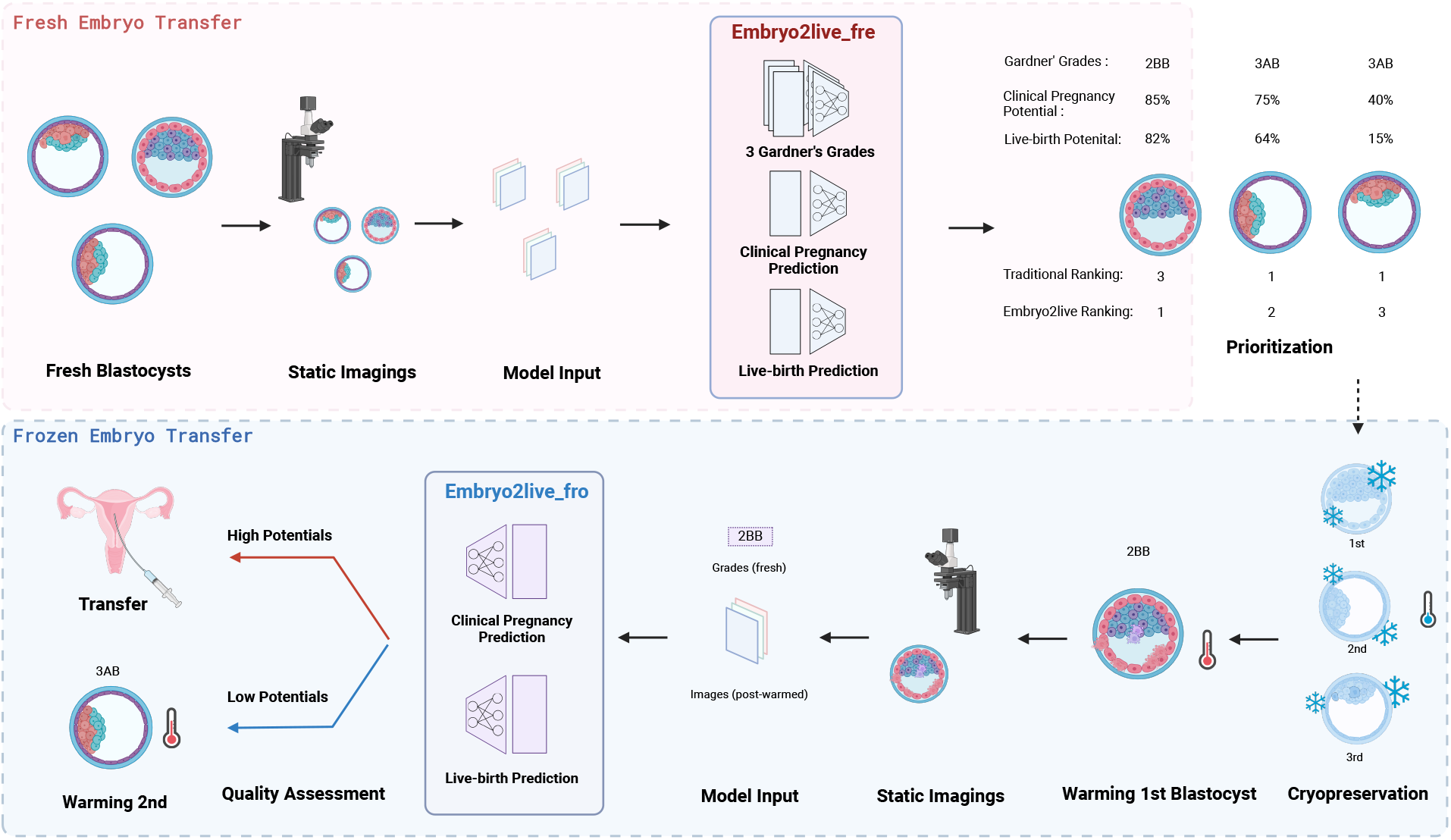
High-level description of the whole framework of Embryo2live. Input the static images of fresh Day5 or Day6 blastocysts in each patient cohort into Embryo2live_fre to get the Gardner’s blastocyst classification grades, clinical pregnancy potential, live birth potential, and also the recommended transfer rank (mainly based on live birth potential) in the fresh embryo transfer. Once frozen embryo transfer is needed, warm the vitrified embryos in rank, and input the fresh numerical grade and post-warmed static image into Embryo2live_fro to check the clinical pregnancy potential and live birth potential after warming. If the embryo quality was below the acceptable level, it would not be suggested to transfer.

Besides fresh transfer, frozen embryo transfer is becoming more common and is particularly beneficial if the uterine conditions of the patients are in a suboptimal state for implantation after ovarian stimulation or there are surplus good quality embryos after transfer (dotted arrow in Fig. 1). In cases of need to proceed to the frozen embryo transfer cycle, one or more vitrified embryos would be warmed in rank by using the static images captured at the fresh stage via the Embryo2live_fre system (Fig. 1 top panel), and the top-ranked embryo would be warmed first. Additionally, our system has a second module Embryo2live_fro to further perform the assessment (or quality check) of the post-warmed embryo by using the image after warming, possibly integrating the morphological grades at the fresh stage.

### 2.2 Embryo2live improves the prediction of Garnder’s classification of embryo grading

To evaluate our Embryo2live_fre system in embryo quality assessment, we started benchmarking the prediction of Gardner’s classification using a recent dataset (n=300 fresh embryos) that has gold-standard annotation, i.e., consensus grades from a consortium of 11 embryologists [9]. The same study has also compiled another dataset with a larger number of samples (n=2,044 fresh embryos) with silver-standard annotations by one senior embryologist. The distributions of the grades on expansion are shown in Fig. 2A, Expansion panel, for our in-house fresh embryo dataset (HKU-QMH CARE EXPANSION), the silver-standard set (B), and the gold-standard set (C). Similarly, the distributions of inner cell mass and trophectoderm are shown in Fig. 2 (Inner Cell Mass, Trophectoderm panels). Of note, for embryos with expansion scores lower than 3 in the dataset, the inner cell mass and trophectoderm scores were omitted by setting not-defined (ND) values.

**Figure 2:**
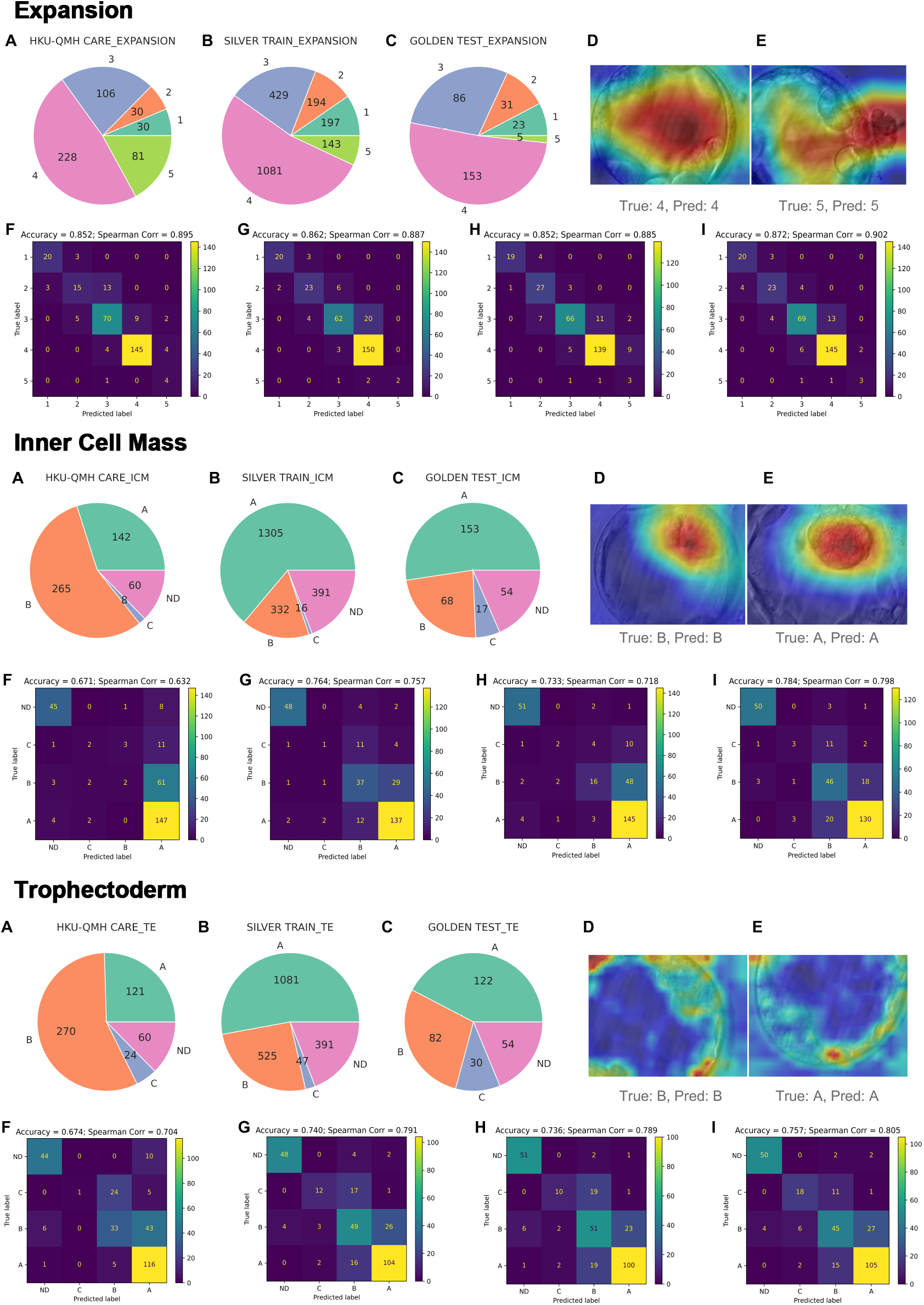
Performance for Gardner’s Grades. Performance for Gardner’s classification in terms of Expansion, Inner cell mass, and Trophectoderm, where Embryo2live outperforms the state-of-the-art methods and supports incorporating external data. A, B, C: Class assignment distribution of Gardner’s criteria within HKU-QMH CARE set (A), the public silver-standard training set (B) and the gold-standard test set (C); D, E: Visual explanations with more attention (red) put on the most relevant region; F, G, H, I: Confusion matrix of the state-of-the-art models (F), Embryo2live model (G), Embryo2live model enhanced by HKU-QMH CARE dataset without label smoothing (H) and Embryo2live model enhanced by HKU-QMH CARE dataset with label smoothing (I).

By using the silver-standard data as the training set and the gold-standard data as the test set, we found that the Embryo2live_fre system consistently outperformed the provided best state-of-the-art models for all three tasks (Fig. 2F, G in Expansion, Inner Cell Mass, Trophectoderm panels). These state-of-the-art models were provided by [9], including XCeption for inner cell mass grade, Swin Transformer for expansion, and trophectoderm grade. Specifically, for predicting expansion, our model further improved the performance given that the state-of-the-art accuracy was satisfactory (accuracy improved from 85.2% to 86.2%). More impressively, the accuracies of inner cell mass and trophectoderm predictions were both increased (from 67.1% to 76.4% for inner cell mass and from 67.4% to 74.0% for trophectoderm). Of note, these three tasks are not independent, particularly considering that the inner cell mass and trophectoderm grades will be marked as ND if the expansion is graded or predicted as lower than 3. Uniquely, our Embryo2live system could output the attention maps (Fig. 2D, E in Expansion, Inner Cell Mass, Trophectoderm panels; see Methods 4.4 on visual explanations), clearly illustrating that our models precisely put most of the weights on the relevant regions. This was particularly remarkable in the trophectoderm prediction: although the captured images failed to show the whole view of the trophectoderm, Embryo2live precisely located and paid more attention to all trophectoderm regions while excluding the inner cell mass region.

Furthermore, we tested if adding our in-house fresh-embryo samples (n=475) into the training set would improve the performance in the above test. By simply combining the silver-standard-annotated training set with our in-house set, we found the prediction on the same gold-standard test set received reduced accuracy for all the 3 tasks (Fig. 2H in Expansion, Inner cell mass, Trophectoderm panels), especially in the inner cell mass task. The decline might be primarily due to differences in grading subjectivity and image-capturing conditions among institutions. As shown in the class assignments (Fig. 2A, B, C in Inner Cell Mass, Trophectoderm panels) the HKU-QMH CARE dataset preferred a conservative estimate of grade B, while the pubic dataset preferred grade A, though the possibility of better embryo quality in the public source cannot be ruled out. To harmonize the annotations, we simply introduced a label smoothing processing in our in-house data with a smoothing factor of 0.2 for expansion and 0.3 for inner cell mass and trophectoderm (See Methods section 4.2 on label preprocessing).

Surprisingly, such smoothing treatment led to a 1% increase in expansion grade accuracy, a 2% increase in inner cell mass accuracy, and a 1.7% increase in trophectoderm accuracy (Fig. 2I in all three panels). We believed that the potential of the label smoothing strategy could be further proven by combining larger datasets and careful adjustment of the annotation scales.

### 2.3 Transfer learning effectively prioritized fresh embryos

Besides predicting Gardner’s classification, our Embryo2live_fre system also supported the prediction of clinical pregnancy and live birth. Here, we used our in-house dataset to evaluate the effectiveness of these tasks. Among our retrospective set of 475 fresh embryos, the success rates of clinical pregnancy and live births per embryo transfer were 35.6% and 32.3%, respectively (Fig. 3B, C, green bar). Of note, we presented the embryologists’ success rate only as a reference instead of a comparison, where the accuracy is taken assuming all the transferred embryos were regarded as live-birth ones.

**Figure 3:**
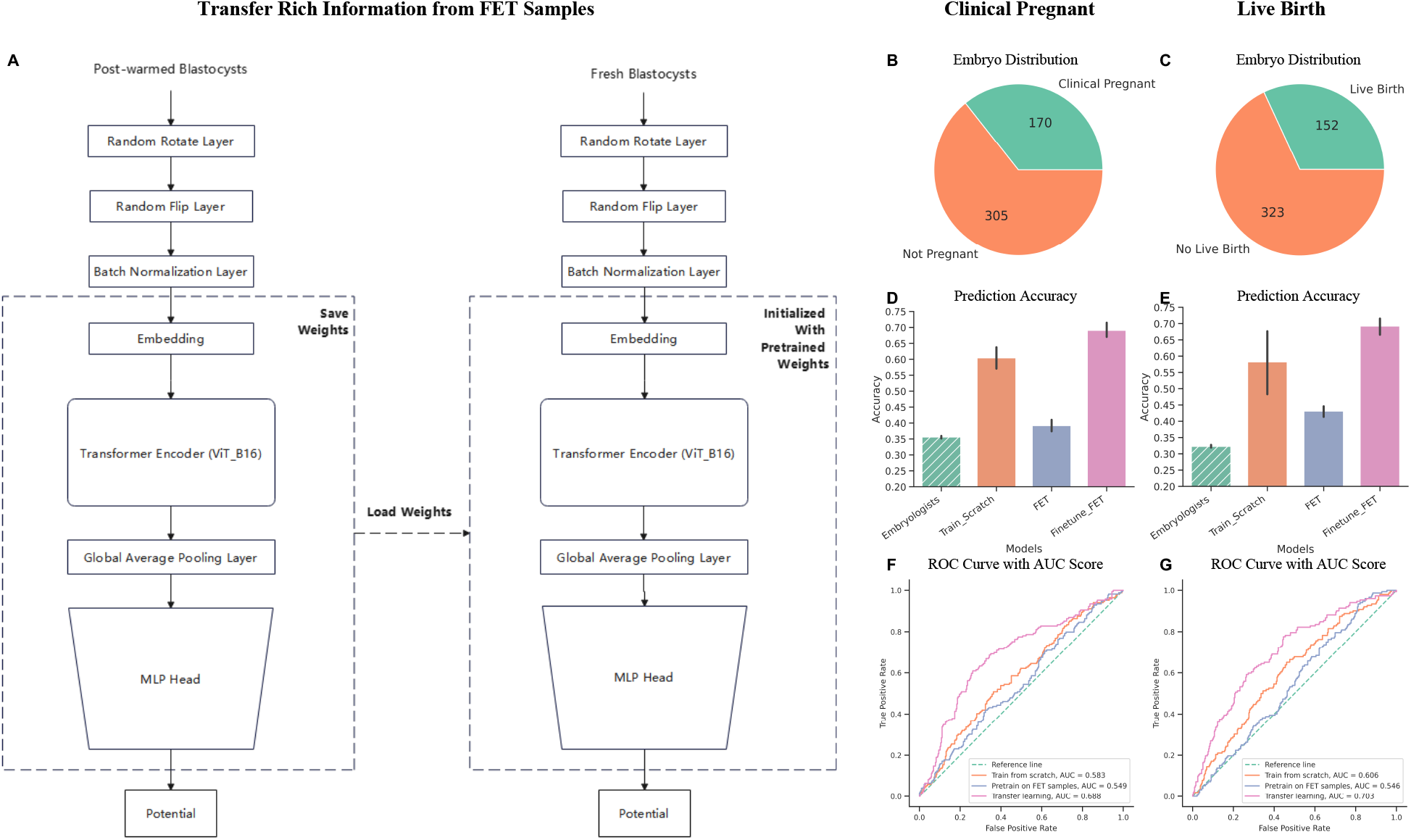
Evaluation of Embryo2live on fresh transfer embryos in predicting clinical pregnancy and live birth. A: The schematic process of transferring rich information from post-warmed embryo to fresh embryo prediction; B, C: Class assignment distribution of the clinical pregnancy (B) and live birth (C) outcome for fresh embryo transfer from HKU-QMH-CARE; D, E: clinical pregnancy (D) and live birth (E) prediction accuracy comparison- clinical records (green bar), train from scratch on fresh samples (orange bar), train on frozen samples without fine-tuning (purple bar), and fine-tuning weights pre-trained on post-warmed samples with small fresh samples (pink bar); F, G: ROC curves and the AUC score for predicting clinical pregnancy (F) and live birth (G) with comparing embryologists as reference (green dash line), training from scratch on fresh samples (orange curve), training on frozen samples without fine-tuning (purple curve), and fine-tuning pre-trained weights on frozen samples with small fresh samples (pink curve).

In contrast, when applying our Embryo2live to the image for the fresh embryos directly, we found obvious improvement compared to the current success rates in both viable pregnancies and live births, with the mean accuracy of 60.4% and 58.2% (Fig. 3D, E orange bar), respectively. This level of accuracy is still moderate, possibly due to the limited data size of fresh embryos.

On the other hand, when performing the same predictions on the frozen embryo transfer dataset, we found our model achieves better performance on both clinical pregnant and live birth predictions (Supp. Fig. S1, purple curve), which may be related to the much larger sample size (n=2,413 embryos). Therefore, we tested if the moderate performance with the small training-set fresh embryos could be improved by adding frozen embryo images and utilizing transfer learning strategies. Specifically, we first pre-trained the model on the post-warmed embryo dataset and fine-tuned it with only 475 fresh embryos using the same 10-fold cross-validation stratification (shown in Fig. 3A). Remarkably, the mean accuracy scores for clinical pregnancy and live-birth prediction increased (from 60.4% and 58.2%) to 69.1% and 69.2%, respectively (Fig. 3D, E; pink bar; with default threshold 0.5). On the contrary, if directly using the trained weights on post-warmed embryos to predict the fresh embryos, it could only get the accuracy of 39.2% and 43.1% (Fig. 3C; purple bar), suggesting the potential attention bias between fresh cycle embryos and post-warmed embryos. Consistently, the ROC curves also demonstrated the effectiveness of the transfer learning with AUC scores reaching 0.688 and 0.703 for clinical pregnancy and live birth prediction respectively (shown in Fig. 3F, G). Overall, with transfer learning by leveraging shared features between fresh and post-warmed embryos, our system resulted in more accurate predictions for both clinical pregnancy and live birth outcomes and proved to be a practically usable tool to assess fresh embryos.

### 2.4 Embryo prioritization reduced time to live birth in patients with multiple embryos available for transfer

As mentioned in Section 2.1, the primary goal of our patient-centered embryo quality assessment is to reduce the time to live birth, especially for patients having multiple cryopreserved blastocysts. In general, this assessment should be conducted at the fresh stage before embryo cryopreservation, while we used the post-warmed image to approximate the rank in the fresh stage given only this stage image is available with known transferred outcomes. Since this is a retrospective dataset, we categorized the patients into five different groups by the number of embryo transfers and their outcomes (Fig. 4A, E in terms of clinical pregnancy and live birth outcomes), which can evaluate the model performance from different perspectives. To train and evaluate the model, we treated the embryos as samples and grouped them by the same patient in a 10-fold cross-validation. Then we reported the performance of individual groups by picking them out from the cross-validation. First, we focused on the single embryo groups (1 and 2) and re-examined the embryo level performance (Supp. Fig. S3), consolidating the consistency when training the model in this setting, i.e., aggregating embryos from all patients despite the uneven embryo cohort sizes (Supp. Fig. S1; Supp. Fig. S2).

**Figure 4:**
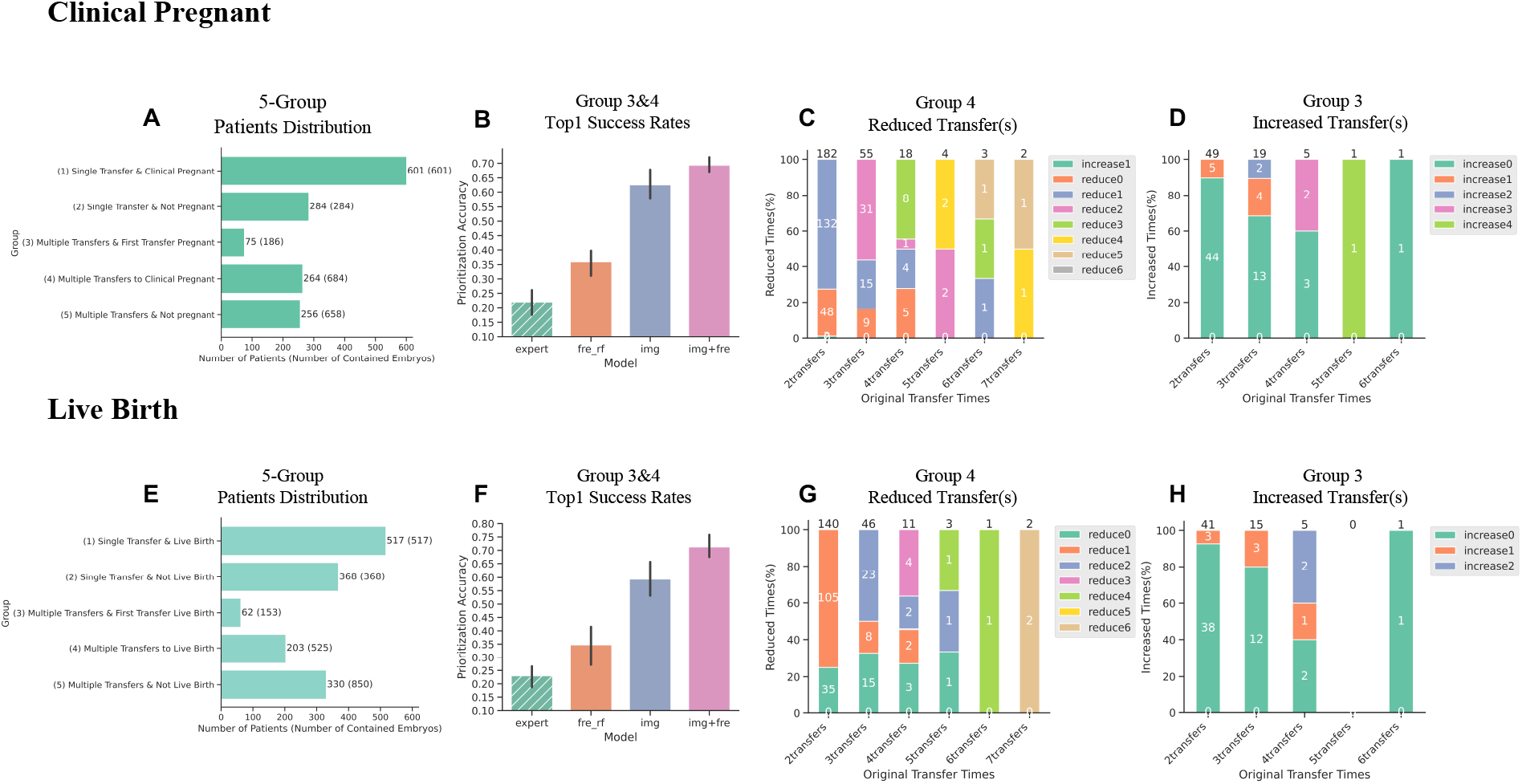
Embryo2live reduces time to clinical pregnancy and live birth, evaluated from the post-warmed embryo dataset. A, E: Patient distributions among five groups according to the number of embryos and outcomes in Clinical Pregnancy (A) and Live Birth (E) settings; B, F: Top1Accuracy within multiple-transfer with clinical pregnancy (B)/ live birth patients (H) in different model settings, expert: embryologists’ first transfer, fre rf: random forest using fresh grade, img: input post-warmed image only, img+fre: input post-warmed image and fresh grade; C, G: Embryo2live reduces the transfer times to clinical pregnancy (C)/ live birth (G) for multiple-transfer to succeed patients (Group 4 as shown in panel A and E); D, H: Embryo2live falsely increases the transfer times to clinical pregnancy (D)/ live birth (H) for multiple-transfer & first transfer succeed patients (Group 3 as shown in panel A and E).

Next, we focused on Groups 3 and 4 which have multiple embryos transfers sequentially and at least one success in clinical pregnancy or live birth in each patient cohort. In particular, we tested if our model can further prioritize the embryos with successful outcomes within each patient, hence reducing the time to live birth. We have excluded Group 5 as there was no clinical pregnancy in the whole cohort. The reason can be multi-factorial, presumably not only due to the embryo quality. From these patients with multiple cryopreserved embryos, our Embryo2live system showed game-changing performance for both clinical pregnancy and live birth (Fig. 4B, F). Specifically, we examined if a certain method could rank the successful embryos as the first place. As a reference, the embryologists only correctly identified 21.9% of clinical pregnancies and 23.0% of live births (i.e., the proportion of Group 3). The random forest classifier can only moderately improve the success rate by using the embryologists’ grades, no matter whether at the fresh or post-warmed stages or combining both (Fig. 4B, F, and Supp. Fig. S4). However, when using the embryo image directly, our Embryo2live system achieves remarkable improvement, for example with top1accuracy of live birth increasing from 46.4% with fresh and cryopreserved grades (fre fro rf) to 71.3% by integrating fresh grades and static images (frozen); Fig. 4B, F). If further diving into Group 4 (i.e., patients with multiple embryos whose success is not the first one) by the number of embryo transfers (2 to 7), we found all groups consistently have more than half of the patients saving over 50% of time, for both clinical pregnancy and live birth (Fig. 4C, G): on average reduced 2.5 to 1.5 transfers for clinical pregnancy and 2.5 to 1.5 transfers for live birth per patient. Most impressively from Fig. 4G, for the two 7-times-transfer patients in Group 4, both could get live birth in the first transfer if following the selection by the Embryo2live system, saving 6 transfers.

On the other hand, we noticed that Group 4 may have a bias on the patient selection that the transfer is likely to stop once a live birth is achieved, limiting the chance to assess if our model may delay the time to live birth. Therefore, we further looked into Group 3 (i.e., patients with multiple embryos cryopreserved and clinical pregnancy or live birth in the first embryo transfer), and we found that our model only makes minimal delay by increasing from 1 transfer to 1.3 for clinical pregnancy (n = 75 patients) and from 1 transfer to 1.2 for live birth (n = 62 patients; Fig. 4D, H). Note, that Group 3 reflected an upper bound of delays that our model might introduce empirically, instead of the expected delays. Taken together, these results suggest that our Embryo2live model can substantially reduce the time to live birth on average (from 2.1 transfer to 1.4, aggregating Groups 3 and 4), with a minimal risk of delay for a certain patient.

## 3 Discussion

Despite that multiple deep learning-based methods have been proposed to enhance embryo assessment, few of them have been deployed in clinics [26], partly due to the lack of a patient-centered setting and the focus on patients with multiple embryos available for transfer. In this study, we introduced the Embryo2live system engineered by advanced deep learning architectures (Vision Transformer and ResNeXt-50) to assess the embryo quality and prioritize the transfer of embryos with higher implantation potential first. By using the public gold-standard dataset, we demonstrated the superior performance of our Embryo2live system in Gardner’s classification, outperforming the state-of-the-art methods by up to 9.3% in accuracy. This system also shows high flexibility to support transfer learning, empowering predicting the clinical pregnancy and live birth of fresh embryos by borrowing the large sample size of cryopreserved embryos.

Most importantly, for the first time, we have demonstrated that AI systems like our Embryo2live can consistently reduce more than half of the embryo transfers in the majority of patients with multiple embryos available for transfer. This improvement is effective for these patients (with a live birth after transfer of ϡ embryos; Group 4 in Fig. 4A), especially for the two 7-embryo-transfer patients both can directly achieve live birth if transferring the embryo selected by our system (reducing 6 failed transfers). With these encouraging results, we are expecting that our Embryo2live system can make a substantial clinical impact.

On the other hand, there are also a few limitations in this study and multiple aspects to further investigate. First, the dataset is retrospective and, therefore, the reduced time to live birth mentioned above may be partly confounded by the sample selection bias. To systematically assess such risk, we evaluated another group of patients with multiple embryos available for transfer (Group 3 in Fig. 4A; success in the first transfer) and found that the empirical upper bound delay was small. Nevertheless, to confirm the effectiveness of the Embryo2live system in embryo selection, a randomized clinical trial is definitely required, before its clinical use.

Second, as demonstrated above, our Embryo2live system is a multivariate prediction system, including Gardner’s grades and potentials of clinical pregnancy and live birth. Hypothetically, when a patient gets pregnant after transferring an embryo with high clinical pregnancy potential but low live birth potential (e.g., Supp. Fig. S5), extra monitoring and care may be offered to the woman after detection of fetal heartbeat at the first ultrasound scan to enhance the chance for a live birth.

Third, in our fresh embryo dataset, we evidenced the power of transfer learning to boost performance when applied to small datasets. Such lack-of-training-set scenarios may broadly exist, particularly in smaller IVF clinics. To resolve this challenge, we have released our pre-trained model in a public repository and allow users to use it for their datasets. Additionally, as a community effort, we anticipate initiatives of federated learning [27] to further support multicenter data sharing and to keep upgrading a community-supported model that can achieve high accuracy and high robustness without releasing patient information.

Last, regarding model backbones, we specifically choose the most suitable architectures based on the inherent nature of the tasks. We employ Convolutional Neural Networks (CNN) for the three grading tasks due to their sensitivity to specific features in local regions. On the other hand, ViT models are used for clinical pregnancy and live-birth prediction tasks, as they can better integrate global features thanks to the self-attention mechanism. But note that in the public dataset, most of the static images lose the full view of the trophectoderm part, so we replace the CNN backbone with ViT for trophectoderm classification, aiming to integrate the global information to make up the defects. From the future perspective, we may anticipate that with a large community-supported dataset, a foundation model may appear soon, to support the multi-task demands, resolving the necessity of training tailored backbone models.

## 4 Methods

### 4.1 Ethical Approval

The study protocol was reviewed and approved by the Institutional Review Boards of the University of Hong Kong/Hospital Authority Hong Kong West Cluster (UW 24-229).

### 4.2 Data Preparation

In this study, we utilized data from two sources. The first source was a public dataset released by Kromp et al. [9], which comprises 2,044 silver-standard Gardner-graded static images (annotated by one senior embryologist) for training (Fig. 2 A2) and 300 gold-standard Gardner-graded images (annotated by 11 embryologists for consensus) for testing (Fig. 2 A3). One limitation of the dataset [9] is that most images primarily focus on the inner cell mass while largely neglecting the trophectoderm. It is worth noting that, in early-stage blastocysts, the inner cell mass and trophectoderm are not yet clearly identifiable, hence these blastocysts are labeled as not defined in this public dataset. Additionally, the dataset contains blastocysts with expansion score from 1 to 5 but not the fully hatched blastocysts (grade 6). We collected the results of state-of-the-art models (Swin Transformer for Expansion prediction, XCeption for inner cell mass prediction, and Swin Transformer for trophectoderm prediction) from the reported Excel files in the GitHub repository: https://github.com/software-competence-center-hagenberg/Blastocyst-Dataset/tree/main/model_predictions.

The second source of data is retrospective information collected from Queen Mary Hospital in Hong Kong (HKU-QMH CARE) between January 2018 and June 2022. This dataset includes 475 static images of transferred Day5 blastocysts in the fresh cycles and 2,413 images of transferred vitrified-warmed blastocysts in frozen embryo transfer cycles, in which clinical pregnancy and live birth outcomes were recorded. For the fresh transfer, embryologists graded the blastocysts based on Gardner’s classification before the transfer. For the frozen embryo transfer cycles, blastocysts underwent two-stage morphological grading: before vitrification in the fresh cycle and after warming (within 2 hours) in the frozen embryo transfer cycle, while the static images were only taken from the post-warm stage. As listed in Fig. 4A and E, the vitrified-warmed embryos came from 1,480 patients. The patients were further grouped into five categories according to the number of transferred embryos and their outcomes:

- Group 1: a single embryo transfer with clinical pregnancy or live birth;
- Group 2: a single embryo transfer without clinical pregnancy or live birth;
- Group 3: multiple embryo transfers with clinical pregnancy or live birth in the first embryo transfer followed by transfer of frozen embryo from the same IVF cycle;
- Group 4: multiple embryo transfers without clinical pregnancy or live birth in the first embryo transfer followed by subsequent transfer with clinical pregnancy or live birth and
- Group 5: multiple embryo transfers in a single IVF cycle without clinical pregnancy and live birth.

We integrated the public training set with the 475 in-house embryo images to train grading models in fresh embryo transfer cycles (class distribution in Fig. 2A). We also employed the in-house embryos for clinical pregnancy and live birth prediction using a 10-fold-stratified cross-validation with each fold sharing the same class ratio (class distribution in Fig. 3B, C). For each training phase, we allocated 12% of the training samples to the validation set. For the cryopreserved embryos, we first adopted a 10-fold cross-validation on the whole set (grouping embryos into folds according to the patient) to train the clinical pregnancy and live birth prediction model with each experiment sharing the same split for a fair evaluation. For each training phase, we allocated 15% of the training samples to the validation set.

### 4.3 Data Preprocessing

#### Image Preprocessing

The public dataset’s images had been pre-cropped the blastocysts from the original images with an aspect ratio of 4:3, thus no further region of interest (ROI) extraction techniques were applied in this work. For the in-house data, the original embryo images contain a large portion of background noise, such as needles and broken cells. Simple edge detection failed to cannily crop the borders due to these distractions. To help the further prediction algorithms focus on the relevant information without being affected by background noise, we manually annotated the embryos in 100 images and used these images to fine-tune a YOLO-V3 model [28] that was pre-trained on ImageNet. This model was based on a mature and efficient object detection algorithm, which we then applied to the entire dataset to crop a new 4:3 image from the original one. The algorithm’s high processing speed also made it suitable for in-time pre-processing in clinics. Considering the limited size of the dataset and the imbalanced class distribution, we applied offline augmentation techniques when training the grading models to enrich and balance the dataset. These techniques included random combinations of CoarseSaltAndPepper, GammaContrast, MotionBlur, GaussianBlur, EnhanceSharpness, Affine, and Embossment by using the imgaug package at https://imgaug.readthedocs.io. We also normalized the pixels of each image to mitigate biases related to microscope optics and lighting conditions and subsequently, resized the image width and height to 256 pixels.

#### Label Preprocessing

We adopted a one-hot encoding strategy for expansion, inner cell mass, and trophectoderm grading classification, with the number of classes equal to 5, 3, and 3, respectively. Given the potential grading bias across different IVF centers or experts due to subjectivity, for example, the public dataset preferred grading to A (Fig. 2A; panels of Inner Cell Mass and Trophectoderm) while in-house grading tended to grade as B (Fig. 2A), integrating labels from the two institutions directly could be problematic. To achieve higher generalization, we introduced a label-smoothing strategy on the in-house dataset. This strategy converts hard labels (i.e., 100%) to 80% and allocated the remaining 20% uncertainty to adjacent classes that lack confidence by embryologists, given a smoothing factor of 0.2. For example, grade B for inner cell mass was encoded as [0, 1, 0] after one-hot encoding, but after label smoothing, the input label was changed to [0.1, 0.8, 0.1]. Grade 1 for expansion will be transformed from [1, 0, 0, 0, 0] to [0.8, 0.2, 0, 0, 0]. This approach reallocated some probabilities from the assigned class to neighboring classes, mitigating overconfidence in potentially mistaken labels due to subjectivity or errors. For clinical pregnancy and live birth prediction, only viable pregnancy and live birth samples was labeled as 1, while any miscarriage cases was considered 0 in the binary classification task.

### 4.4 Model Development and Training Strategies

The pre-processed 3-channel input images first underwent online augmentation layers, such as horizontal and vertical flip, and rotation, to further improve data diversity and enhance robustness. The images were then proceeded through a batch normalization layer, deep feature extraction layers, and multi-layer perceptron (MLP) layers. The feature extraction layer model repository included ImageNet-pretrained CNN-based models such as Xception, a series of ResNet, DenseNet, EfficientNet, and ViT variants. We adopted an Adam optimizer with a mini-batch size of 32 and a learning rate decay strategy to accelerate convergence in the model optimization. The maximum training epoch was set to 80, with early stopping applied to prevent over-fitting. Specifically, as an imbalance could lead to biased model predictions, where the majority class dominated the learning process, and the minority class was often overlooked, for clinical pregnancy and live birth prediction cases, we computed and assigned class weights based on input label distribution to penalize false predictions in the minority class. We trained each model using a single NVIDIA A100 GPU, with all training processes implemented on TensorFlow-GPU v2.6.0.

For grading, we aimed to match the most suitable model backbones for different grading tasks. The final model backbone for expansion grade prediction was ResNext-50 [24] with three MLP layers after the GlobalAveragePooling layer (dimensions: 1000, 512, 5; activations: softmax, tanh, softmax, respectively). For inner cell mass grade prediction, we still used ResNext-50 while two MLP layers were added after it (dimensions: 1000, 3; activations: softmax, softmax, respectively). For trophectoderm grade prediction, we selected ViT-b16 [25] with two MLP layers (dimensions: 1000, 3; activations: softmax, softmax) after the GlobalAveragePooling layer. For the ultimate output Gardner’s grade, if the expansion grade was predicted to be lower than 3, the inner cell mass and trophectoderm grades would be automatically changed to not defined no matter what the original prediction was.

As there were limited fresh cycle embryos used in this work for clinical pregnancy and live birth prediction but more cryopreserved embryos were collected, we adopted transfer learning a method where a model developed for one task could be reused as the starting point for another related task. The fresh and the vitrified-warmed embryos undoubtedly shared many common features, therefore we transferred this rich information by pre-training on frozen embryos and using fresh embryos data to fine-tune the model. The final model backbone design for these tasks used ViT-b16 for feature extraction, followed by three MLP layers after the GlobalAveragePooling layer in pregnant prediction (dimensions: 1000, 64, 1; activations: softmax, tanh, sigmoid) and by four layers in live birth prediction (dimensions: 1000, 256, 64, 1; activations: softmax, elu, tanh, sigmoid).

For models used in frozen embryo transfer cycles, one model design inputted only images (post-warmed), while the other design inputted both the post-warmed static images and the numerical grade at its fresh stage. The two settings were tested under the same model backbone and initial hyper-parameters. The model backbone chosen for these tasks is ViT-b16, combined with two MLP layers after the GlobalAveragePooling layer (dimensions: 1000, 196; activations: softmax, elu), one dropout layer (dropout rate=0.3), and two additional MLP layers (dimensions: 64, 1; activations: tanh, sigmoid). We specifically assigned the larger model ViT-l16 in live birth prediction with the same MLP heads setting. Note, that all the contrast models used in this work shared the same random seed, data split, model backbone, hyperparameters, and earlystopping strategies.

### 4.5 Evaluation

#### Loss Functions

Considering that classes of Expansion, inner cell mass, and trophectoderm were not independent as there was an inherent order between the classes, but not a meaningful numeric difference between them, and thus the loss function for the grading model should take into account the degree of misclassification. For instance, when predicting the expansion grades, if the true label is 2, a false prediction of 5 should be penalized more than a false prediction of 3. Therefore, the loss function for the grading model was formulated as:

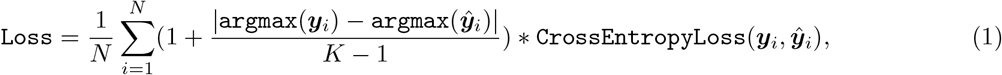

where *K* represents the number of classes in each classification task, and *N* denotes the number of samples, *y*_*i*_ and *ŷ*_*i*_ denotes the observed and predicted label probability vectors, respectively. This loss function ensured that predictions further from the true label were penalized more heavily, providing a more prudent evaluation of the model’s performance in ordinal classification scenarios so that even false predictions were still close to the true grade. For clinical pregnancy and live birth predictions, we simply adopted the binary cross entropy loss function with assigned class weights.

#### Evaluation Metrics

For the grading classification task, we employed accuracy in terms of each embryo as the evaluation metric to assess the classification performance, that y_pred equaled the class with the highest probability after softmax activation. For the clinical pregnancy and live birth models used in fresh transfer samples, we compared the prediction accuracy score with embryologists’ success rates with the default threshold (0.5) and also the ROC curves with AUC scores.

For the clinical pregnancy and live birth models used in frozen embryo transfer cycles, we judged the performance in terms of patient cohorts rather than embryos. The aim was to ensure a more patient-centric evaluation of the model’s performance in identifying the most viable embryos for each individual, better reflecting the clinical application. To be specific, we aimed to validate the ability to prioritize to transfer the embryo with the highest implantation potential in a patient cohort, by calculating the Top1 Accuracy (Top1 recall rate):

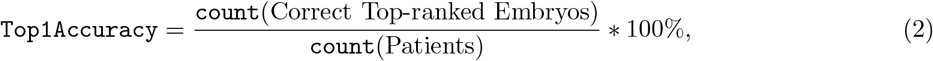

where, the top-ranked embryo is the embryo with the highest predicted potential in a patient cohort and it would be considered as correct if it matches with the final pregnant or live-birth one. The denominator refers to the total number of patients (in Group 3&4) who have multiple transfers in an IVF cycle and achieve at least one clinical pregnancy or live-birth outcome.

#### Visual explanations

In the Gardner’s classification tasks, we introduced visual explanation techniques to help understand how ‘black box’ deep learning models arrived at their output from embryo imaging data [29], potentially making the learning more explicit and more interpretable. In expansion and inner cell mass predictions (CNN-based models), the Gradient-weighted Class Activation Mapping algorithm (Grad-CAM) [30] was applied to show the class activation maps for each embryo by flowing into the final convolutional layer to produce a coarse localization map highlighting the important regions in the image. In trophectoderm classification (ViT-based models), we used Attention Rollout [31] to visualize the attention maps for each embryo by averaging attention weights across all heads and then recursively multiplying the weight matrices of all layers [25].

## Data Availability

For the data collected by HKU-QMH-CARE, data sharing will be granted by reasonable request.

https://doi.org/10.6084/m9.figshare.25479469.v1

https://doi.org/10.6084/m9.figshare.20123153.v3

## Data availability

The dataset for assessing grading prediction is produced by [9], where all images are downloaded from the figureshare repository at https://doi.org/10.6084/m9.figshare.20123153.v3. For the data collected by HKU-QMH-CARE, data sharing will be granted by reasonable request.

## Codes availability

The source codes of Embryo2live are all freely available at https://github.com/louisa9634/Embryo2live. Detailed workflows to reproduce figures and results in this paper were written as Jupyter notebooks in the repository. The pre-trained models from all in-house frozen embryos and YOLO detection models are available on a figureshare repository at https://doi.org/10.6084/m9.figshare.25479469.v1.

## Acknowledgments

The authors would like to thank all the embryologists in HKU-QMH CARE for their technical support. This project is supported by the National Natural Science Foundation of China (No. 62222217), Innovation Technology Commission Funding (Health@InnoHK), the University of Hong Kong through a startup fund and a seed fund (Y.H.), Shenzhen Fundamental Research Program of China (No. JCYJ20220818103013028), Shenzhen Science and Technology Program (KQTD20190929172749226), and Shenzhen Sanming Project of Medicine (SZSM202211014).

## Author Contributions

E.N. and W.Y. conceived the project. Y.L.L. and Y.H. supervised the study. Lu Yu implemented the Embryo2live models and performed all analyses with support from Y.H. and Lequan Yu. K.L. performed embryo grading and image generation for all in-house data. Lu Yu, Y.H., and Y.L.L. wrote the manuscript with inputs from all authors.

