## supplementary for "Deep learning-based embryo assessment of static images can reduce the time to live birth in *in vitro* fertilization"

**Additional file 1**

Figure S1. ROC-AUC comparisons on different model settings in frozen embryo transfer prediction among all embryo samples.
Figure S2. Accuracy comparisons on different model settings in frozen embryo transfer prediction among all embryo samples
Figure S3. Embryo2live: Validate Embryo2live by ROC curve (orange) with AUC score for single-transfer patients.
Figure S4. Top1Accuracy within multiple-transfer with clinical pregnancy (left)/ live birth patients (right) in different model settings.
Figure S5. Relate Clinical Pregnancy Potentials to Live Birth Potentials.
Table S1. Group 4 reduced transfer(s) to clinical pregnancy.
Table S2. Group 3 increased transfer(s) to clinical pregnancy.
Table S3. Group 4 reduced transfer(s) to live birth.
Table S4. Group 3 increased transfer(s) to live birth.

### ROC-AUC Comparisons of Different Model Settings in FET Predictions

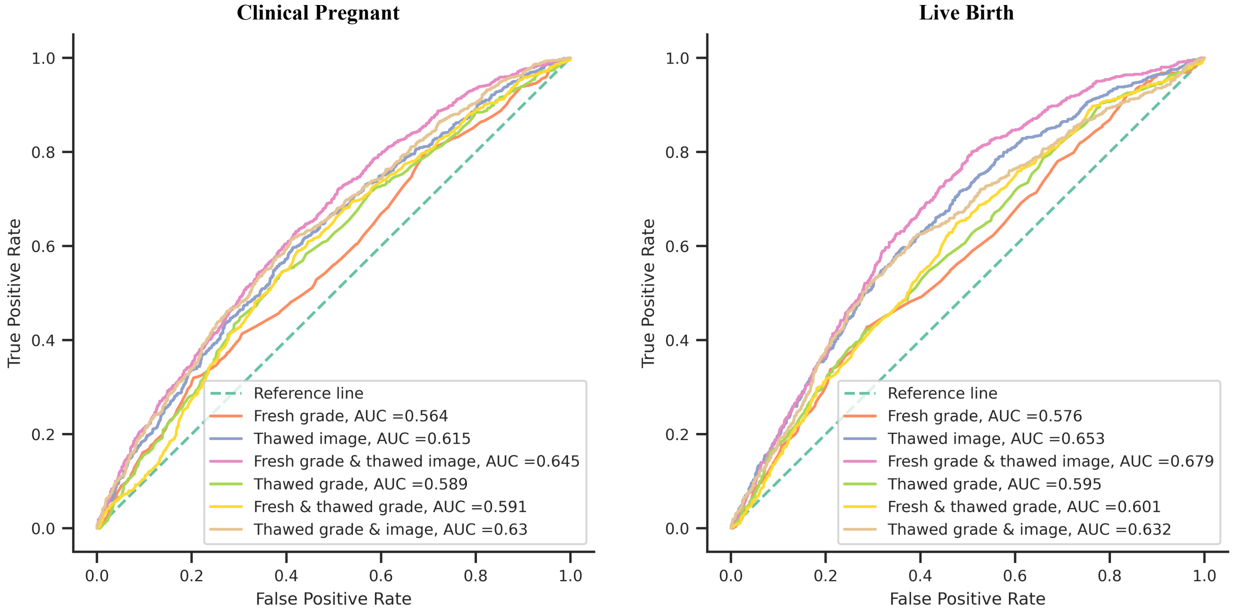


**Figure** S1: ROC-AUC comparisons on different model settings in frozen embryo transfer prediction among all embryo samples; Reference line: embryologists' first transfer, Fresh grade: random forest using fresh grade; Thawed image: input post-warmed image only, input post-warmed image with fresh grade; Fresh grade & thawed image: input post-warmed image with fresh grade; Thawed grade: random forest using frozen grade; Fresh & thawed grade: random forest using fresh and frozen numerical grade; Thawed grade & fresh image: input post-warmed image and grade.

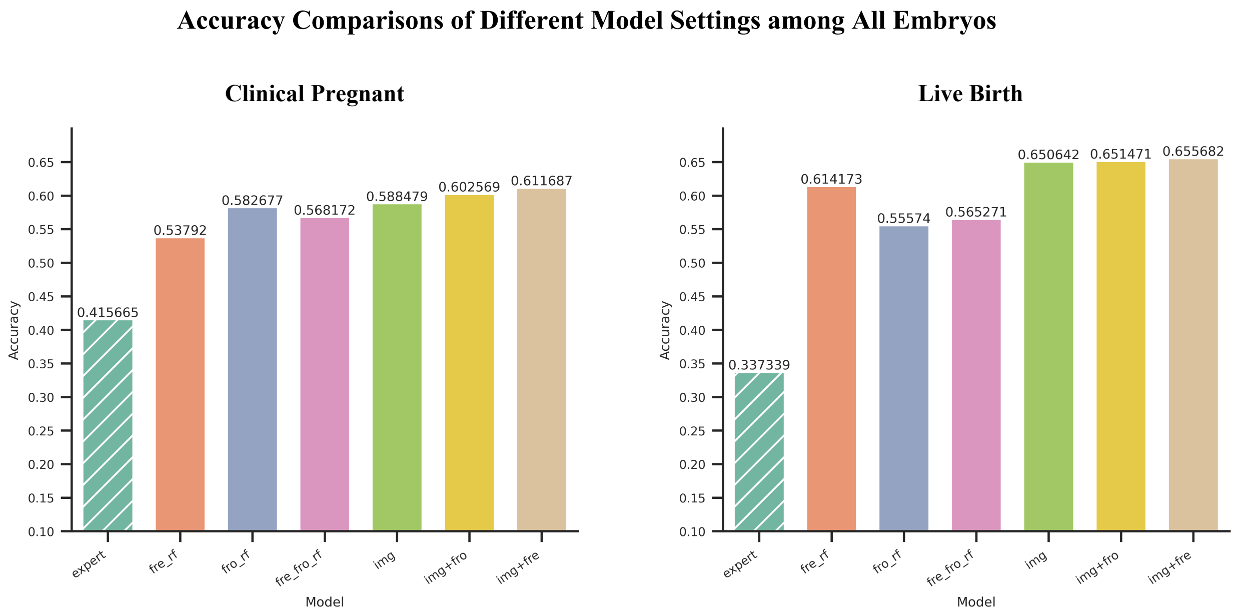


**Figure** S2: Accuracy comparisons on different model settings in frozen embryo transfer prediction among all embryo samples; expert: embryologists' first transfer, fre_rf: random forest using fresh grade; fro_rf: random forest using frozen grade; fre_fro_rf: random forest using fresh and frozen numerical grade; img: input post-warmed image only; img+fro: input post-warmed image and grade; img+fre: input post-warmed image with fresh grade.

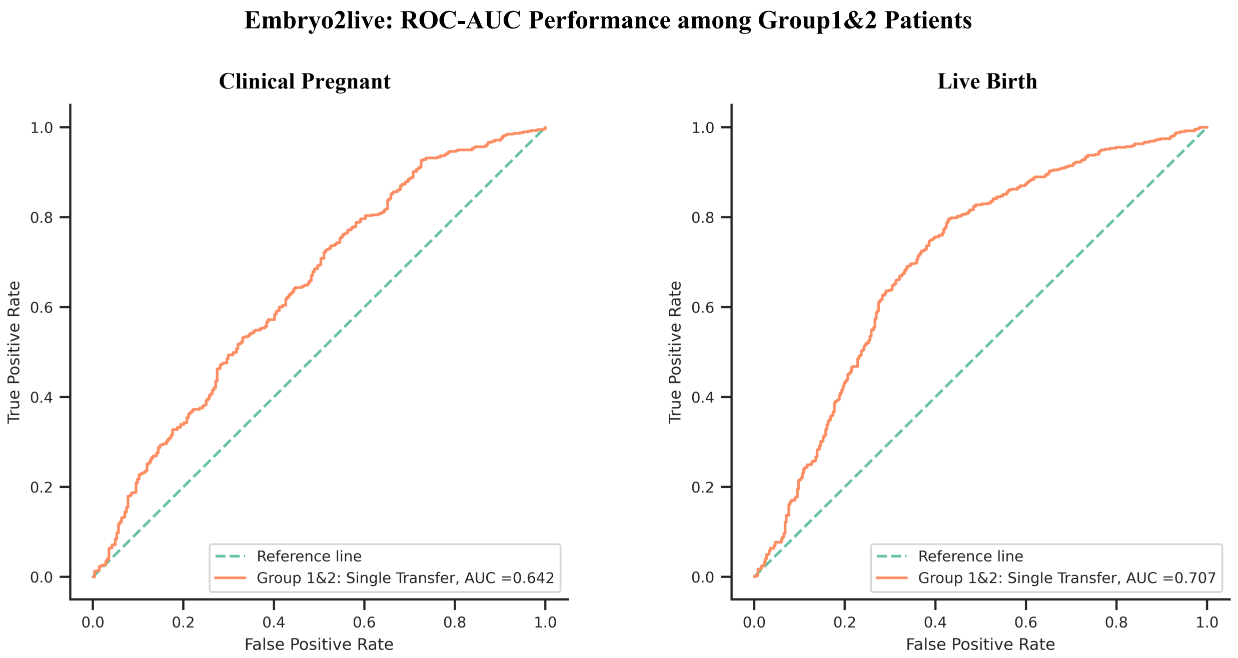


**Figure** S3: Embryo2live: Validate Embryo2live by ROC curve (orange) with AUC score for single-transfer patients

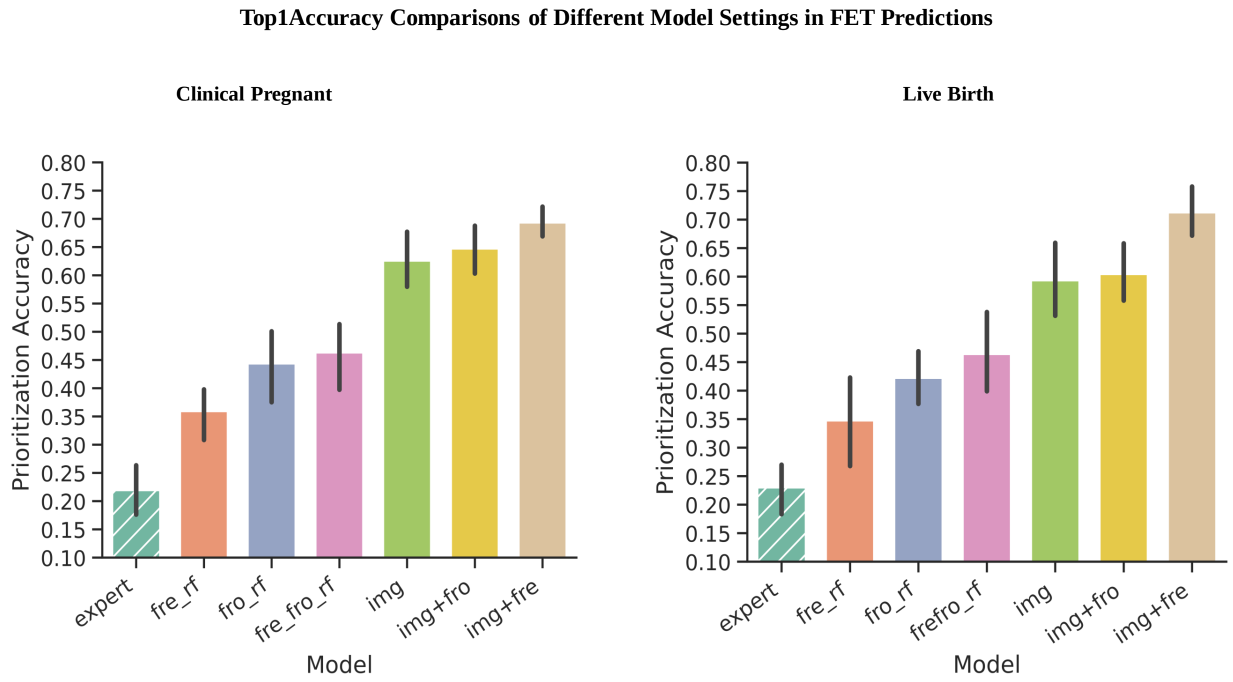


**Figure** S4: Top1Accuracy within multiple-transfer with clinical pregnancy (left)/ live birth patients (right) in different model settings; expert: embryologists' first transfer, fre_rf: random forest using fresh grade, fro_rf: random forest using frozen grade, fre_fro_rf: random forest using fresh and frozen numerical grades, img: input post-warmed image only, img+fro: input post-warmed image and grade, img+fre: input post-warmed image with fresh grade.

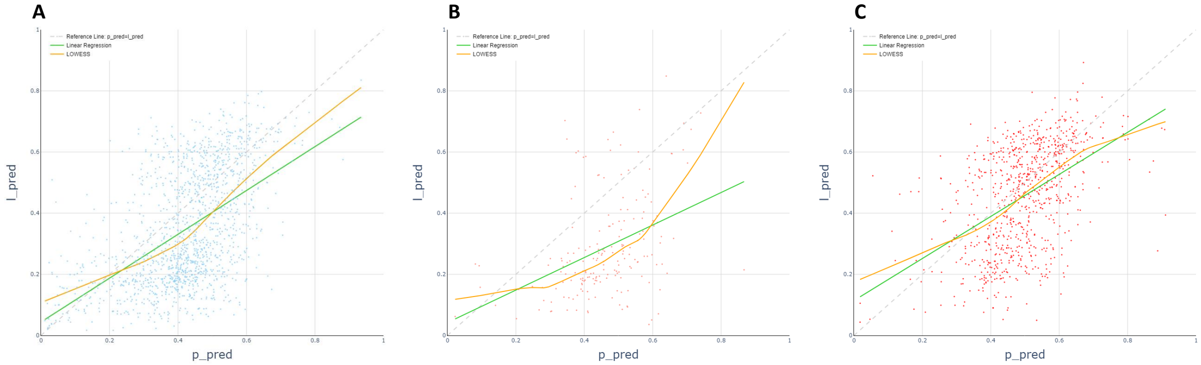


**Figure** S5: Relate Clinical Pregnancy Potentials to Live Birth Potentials for (A): Not Clinical Pregnancy, (B) Clinical Pregnancy but No Live Birth, and (C) Live Birth Embryo, fitted by linear regression (green) and local regression (LOWESS, orange), dashed lightgrey diagnoal line as reference.

Table 1: Group 4 reduced transfer(s) to clinical pregnancy

| | increase1 | reduce0 | reduce1 | reduce2 | reduce3 | reduce4 | reduce5 | reduce6 |
|---|---|---|---|---|---|---|---|---|
| **2transfers** | 2 | 48 | 132 | 0 | 0 | 0 | 0 | 0 |
| **3transfers** | 0 | 9 | 15 | 31 | 0 | 0 | 0 | 0 |
| **4transfers** | 0 | 5 | 4 | 1 | 8 | 0 | 0 | 0 |
| **5transfers** | 0 | 0 | 0 | 2 | 0 | 2 | 0 | 0 |
| **6transfers** | 0 | 0 | 1 | 0 | 1 | 0 | 1 | 0 |
| **7transfers** | 0 | 0 | 0 | 0 | 0 | 1 | 1 | 0 |

Table 2: Group 3 increased transfer(s) to clinical pregnancy

| | increase0 | increase1 | increase2 | increase3 | increase4 |
|---|---|---|---|---|---|
| **2transfers** | 44 | 5 | 0 | 0 | 0 |
| **3transfers** | 13 | 4 | 2 | 0 | 0 |
| **4transfers** | 3 | 0 | 0 | 2 | 0 |
| **5transfers** | 0 | 0 | 0 | 0 | 1 |
| **6transfers** | 1 | 0 | 0 | 0 | 0 |

Table 3: Group 4 reduced transfer(s) to live birth

| | reduce0 | reduce1 | reduce2 | reduce3 | reduce4 | reduce5 | reduce6 |
|---|---|---|---|---|---|---|---|
| **2transfers** | 35 | 105 | 0 | 0 | 0 | 0 | 0 |
| **3transfers** | 15 | 8 | 23 | 0 | 0 | 0 | 0 |
| **4transfers** | 3 | 2 | 2 | 4 | 0 | 0 | 0 |
| **5transfers** | 1 | 0 | 1 | 0 | 1 | 0 | 0 |
| **6transfers** | 0 | 0 | 0 | 0 | 1 | 0 | 0 |
| **7transfers** | 0 | 0 | 0 | 0 | 0 | 0 | 2 |

Table 4: Group 3 increased transfer(s) to live birth

| | increase0 | increase1 | increase2 |
|---|---|---|---|
| **2transfers** | 38 | 3 | 0 |
| **3transfers** | 12 | 3 | 0 |
| **4transfers** | 2 | 1 | 2 |
| **5transfers** | 0 | 0 | 0 |
| **6transfers** | 1 | 0 | 0 |
